# Review of published systematic reviews and meta-analyses on COVID-19

**DOI:** 10.1101/2020.06.03.20121137

**Authors:** Nasrien E. Ibrahim, Ezzeldin M. Ibrahim

## Abstract

**Purpose:** The rapid spread of the COVID-19 pandemic has prompted researchers from all over the world to share their experience. The results were numerous reports with variable quality. The latter has provided an impetus to examine all published meta-analyses and systematic reviews on COVID-19 to date to examine available evidence. Methods: Using predefined selection criteria, a literature search identified 43 eligible meta-analyses and/or systematic reviews. Results: Most (N=17) studies addressed clinical manifestations and associated comorbidity, 6 studies addressed clinical manifestations in pregnant women and younger individuals, 8 studies addressed diagnostic data, 9 studies addressed various interventions, and 9 studies addressed prevention and control. The number of studies included in the various systemic reviews and meta-analyses ranged from 2 to 89. While there were some similarities and consistency for some findings, e.g. the relation between comorbidities and disease severity, we also noted occasionally conflicting data. Conclusion: As more data are collected from patients infected with COVID-19 all over the world, more studies will undoubtedly be published and attention to scientific accuracy in the performance of trials must be exercised to inform clinical decision-making and treatment guidelines.

## Introduction

In December 2019, an outbreak of a new infectious disease in Wuhan in the Hubei Province of China was announced (*1*). Little did we know; this disease would change the world. The disease was caused by a beta-coronavirus called severe acute respiratory syndrome coronavirus 2 (SARS-CoV-2), and the disease was recently named coronavirus disease 2019 (COVID-19). On March 11, 2020, when there were around 120,000 confirmed COVID-19 cases in over 10 countries, the World Health Organization declared COVID-19 a pandemic. As of 30 April 2020, 3,250,267 cases have been reported across 185 countries, resulting in 234,701 deaths, and 1,052,550 people have recovered (*2*).

The rapid spread of the COVID-19 pandemic, its alarming severity and mortality, and the resultant detrimental impact on world economy and social and financial needs of millions of people across the globe, have prompted researchers and clinicians from all over the world to share their experience. For better or for worse, a mass of scientific research has been published, as COVID-19 has created the need for fast dissemination of information about the pandemic. Published and preprint manuscripts are shared widely via news and social media outlets, allowing the experiences to be shared across the globe. Nevertheless, there has been recent concern about the quality of COVID-19 research, and how rigorous the peer review processes were in this era of rapid dissemination (*3, 4*). While the experiences should be shared rapidly and broadly since this is a novel disease process, care must be taken to distinguish between anecdotal experiences and data from randomized controlled trials- the standard clinical trials based on which treatment guidelines are usually created.

In this review, we examined all published meta-analyses and systematic reviews about COVID-19 to date. While we did not intent to evaluate the quality of published studies, we planned to thoroughly examine the data provided and we attempted to compare studies that addressed similar research questions to establish consistency or discrepancy between reports.

## Methods

### Search strategy

Between January 1st, 2020 and April 30th, 2019, we identified eligible studies using electronic literature search of the following databases: MEDLINE, EMBASE, and the Cochrane Library. We used Medical Subject Heading terms or keywords using a combination of the following search terms: “2019 novel coronavirus,” OR “COVID-19,” OR “Coronavirus,” OR “SARS-CoV-2,” OR “Chinese Coronavirus”. The search terms were combined with the publication types “systematic review,” OR “meta-analysis”. Following screening of retrieved records for the relevant titles, the relevant abstracts were reviewed, subsequently, the full text articles were obtained to determine appropriateness for final inclusion.

### Selection criteria

We included all studies that met the following criteria: (1) published in English language between January 1^st^, 2020 and April 30^th^, 2020; (2) only published as systematic review, or meta-analysis, or both; (3) examining any of the following: clinical features, diagnosis, associated comorbidity, treatment and intervention, or prevention and control of COVID-19. Studies that addressed any combination of those features were also considered eligible; and (4) including relevant data for any age, gender, race, or a specific risk group. We also intended to include duplicate articles if they provided additional relevant data.

### Data extraction

All authors reviewed the full text of potential articles and discussed the data intended for extraction and decisions were documented. Extracted data included the following fields: first author last name, the purpose of the study, the type of the study being a systematic review; a meta-analysis; or both, number of included articles, number of included patients, pertinent cross references, and the relevant findings.

## Results

We identified 7,330 potentially relevant articles. After exclusion of duplicate references, non-relevant literature, and those that did not satisfy the inclusion criteria, 43 systematic reviews/meta-analyses were included (*5-47*).

**Table 1** depicts the summary of the 43 included systematic reviews/meta-analyses. Most (N=17) studies addressed clinical manifestations and associated comorbidity, 6 studies addressed clinical manifestations in pregnant women and younger individuals, 8 studies addressed diagnostic data, 9 studies addressed various interventions, and 9 studies addressed prevention and control. Several studies were designed to report on more than one aspect. The number of studies included in the various systemic reviews and meta-analyses ranged from 2 to 89 and some systemic reviews and meta-analyses included no COVID trials. In Table 2 highlights the most significant findings derived from each systematic review/meta-analysis classified according to the research objective(s).

**Table 1.**
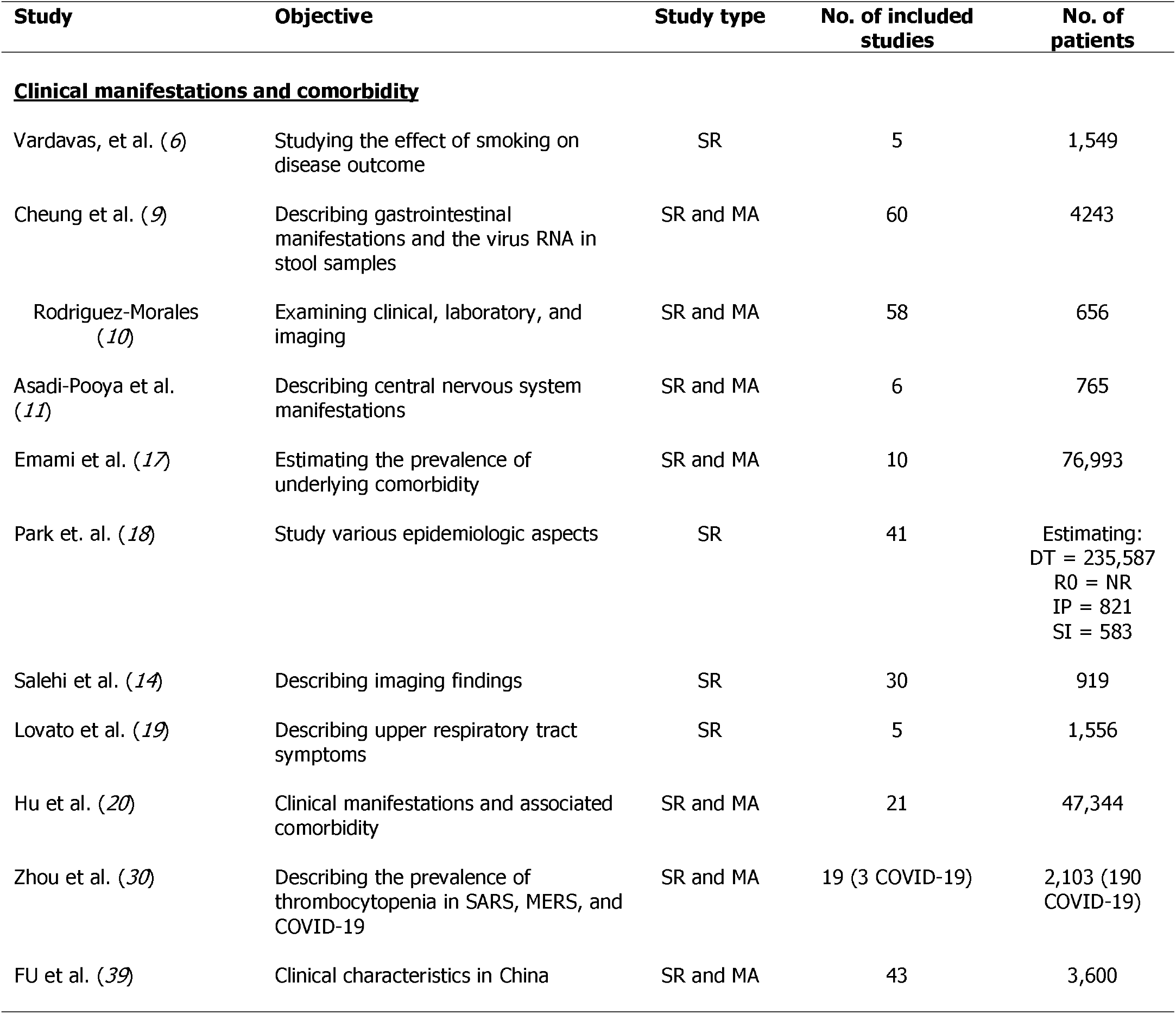
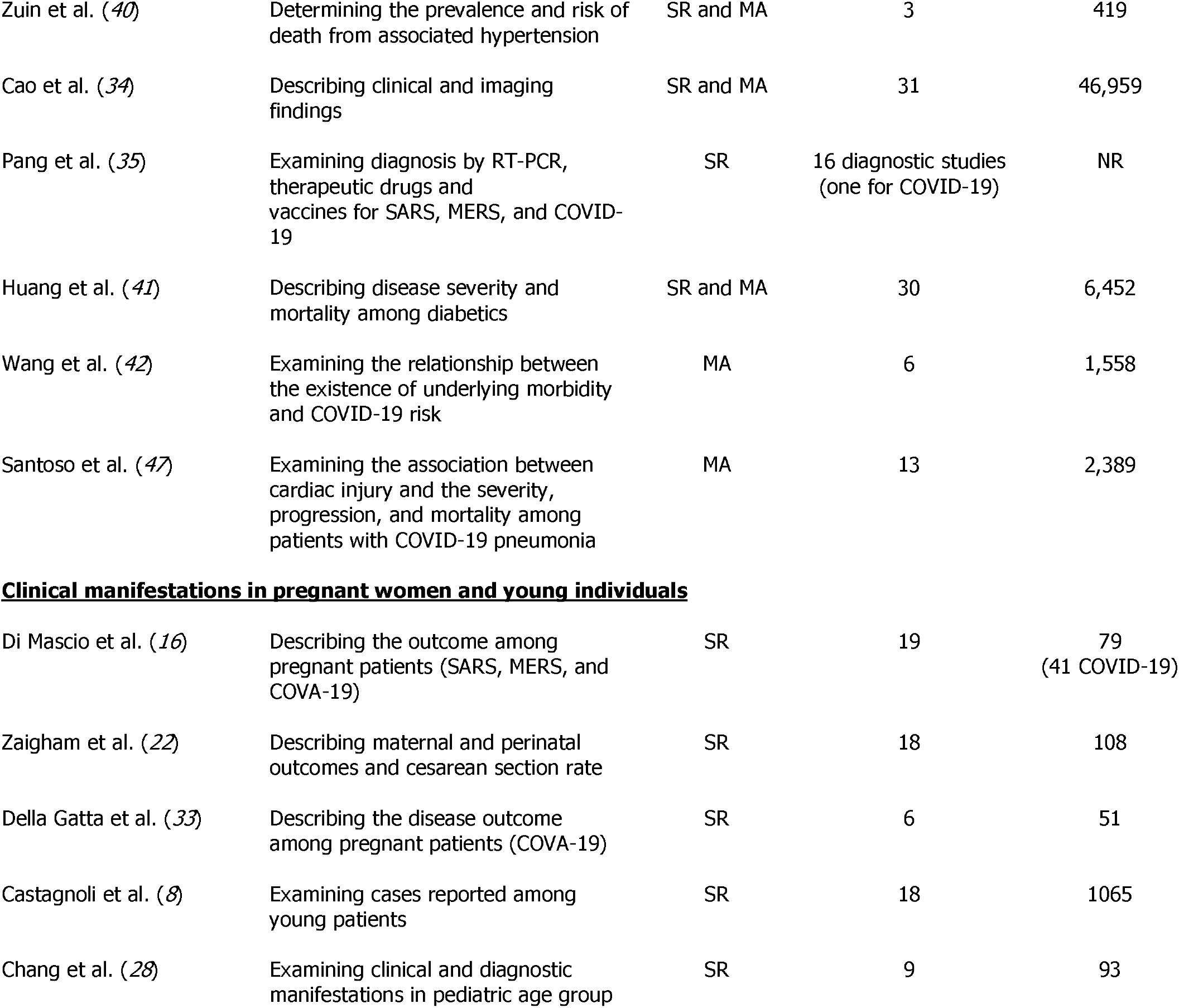
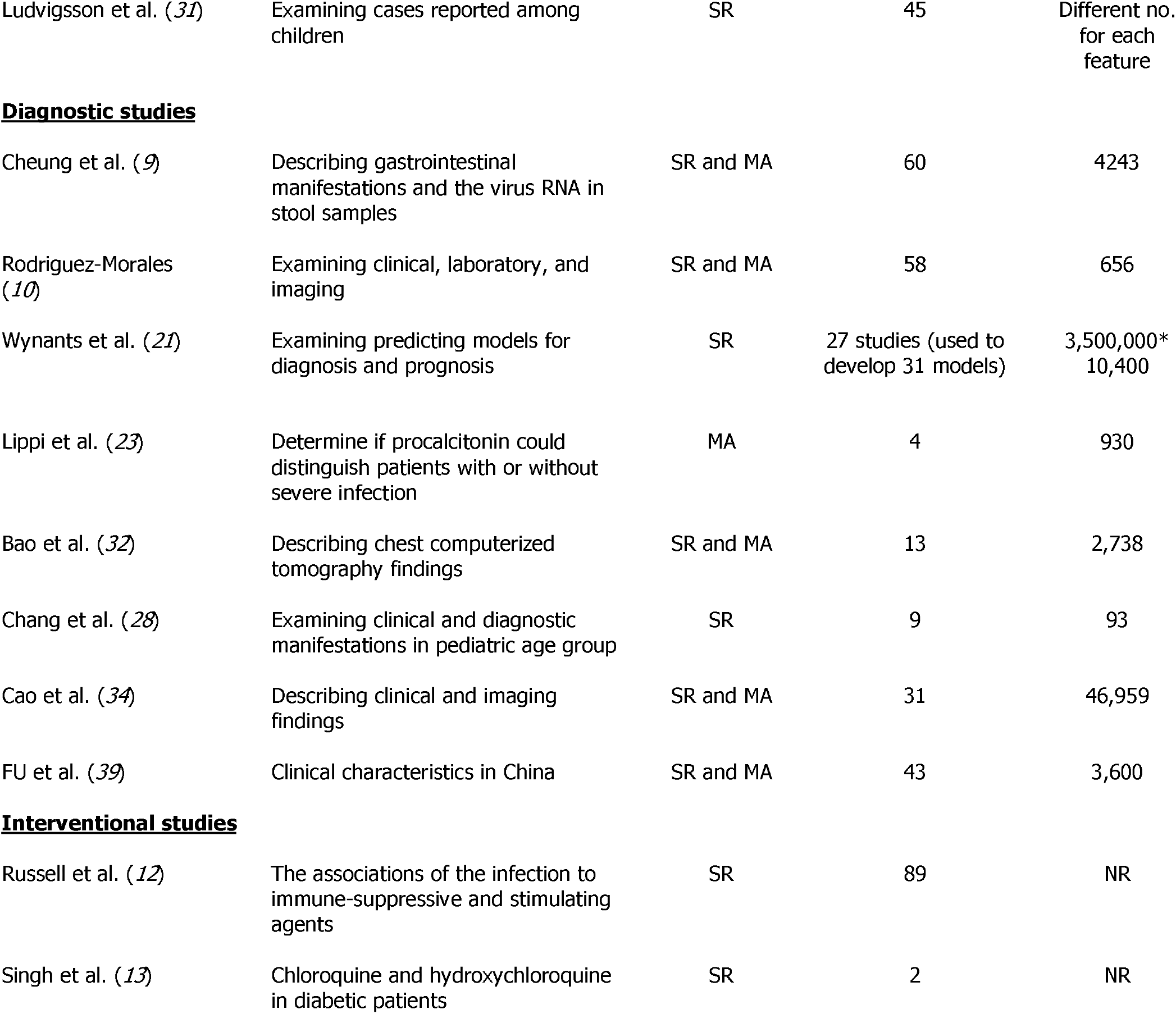
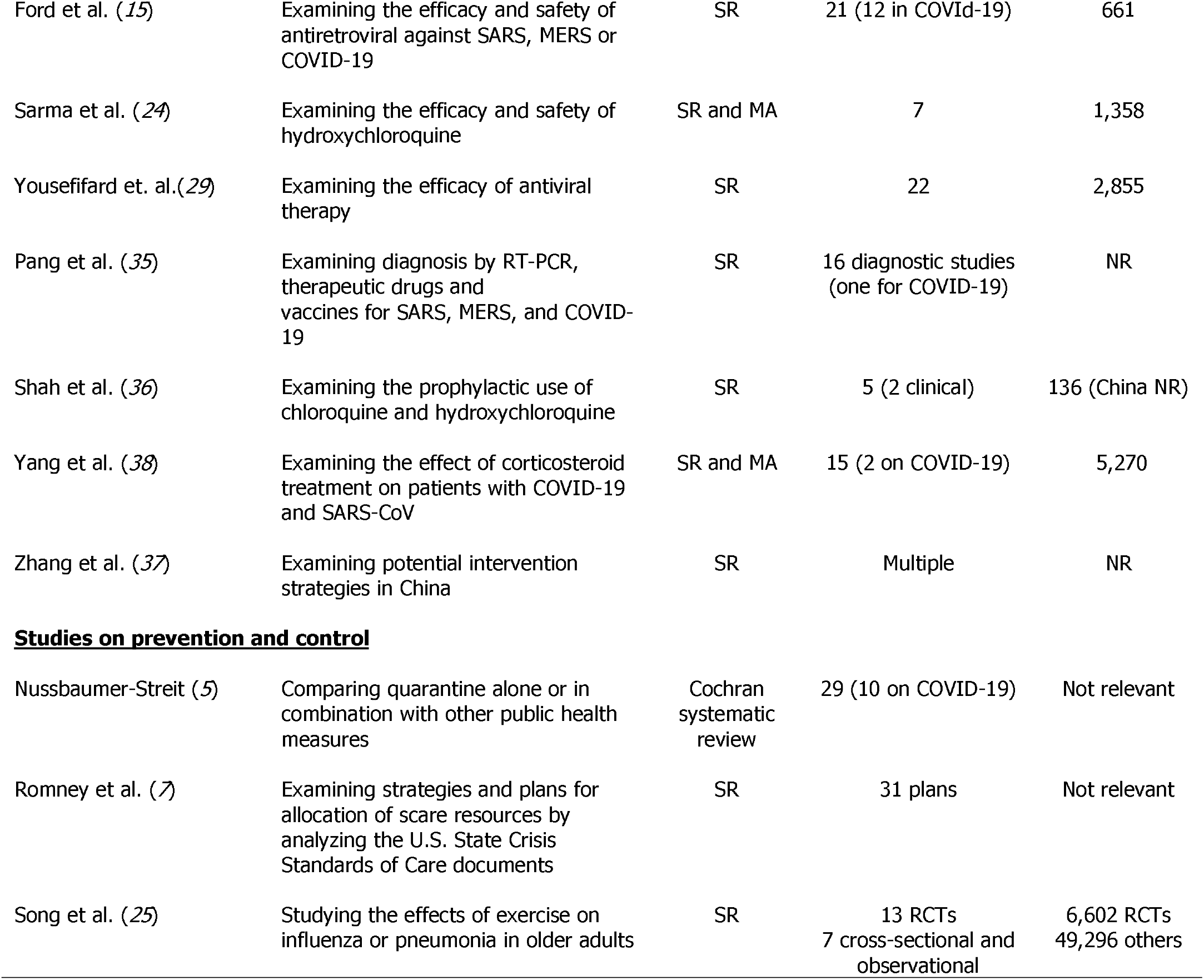
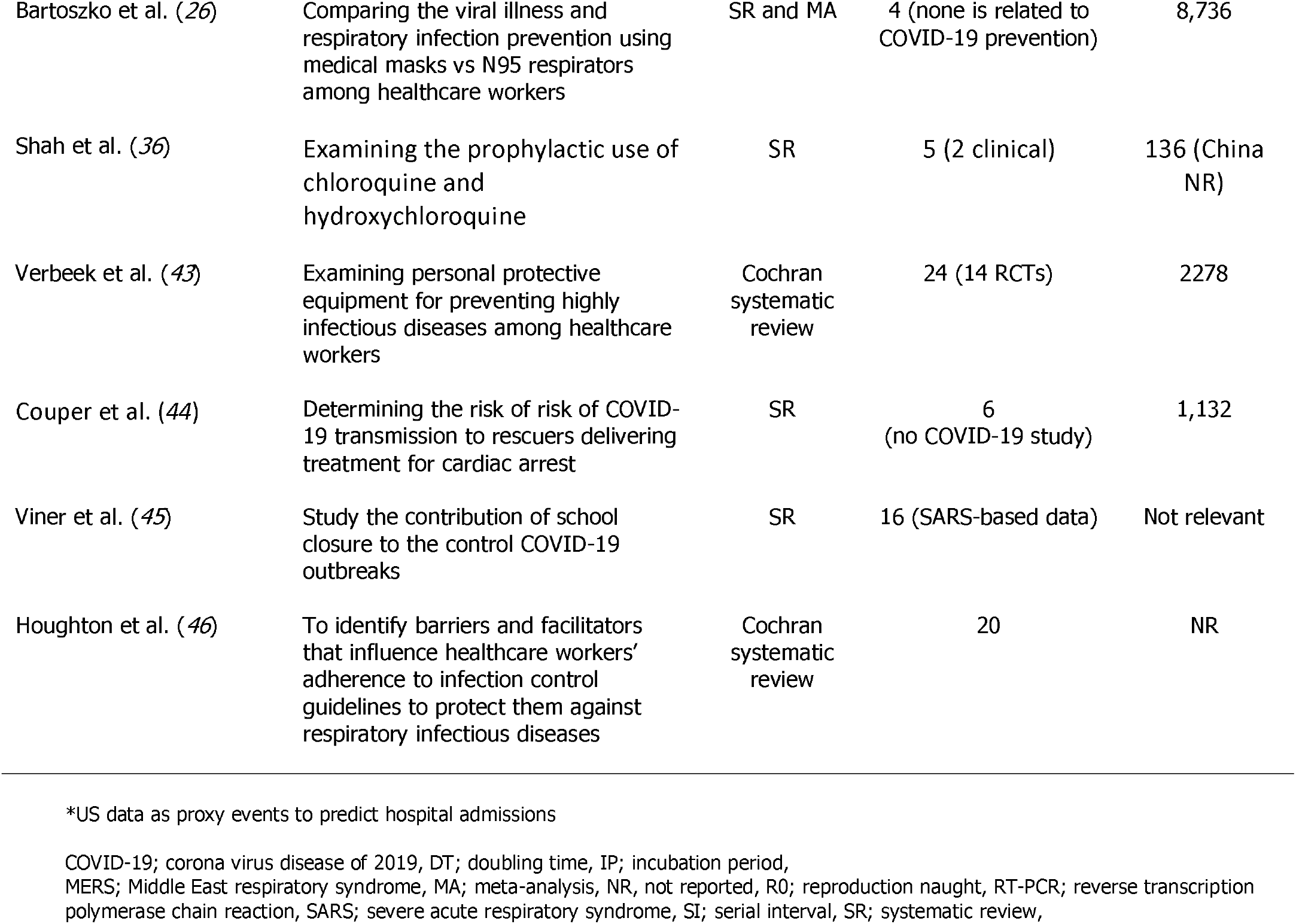
Summary of the objectives and designs of the 43 included studies.

**Table 2** highlights the most significant findings derived from each systematic review/meta-analysis classified according to the research objective(s). The systematic reviews/meta-analyses we reviewed included studies with occasionally disparate outcomes. The systematic review by Vardavas et al (*6*) included studies that demonstrated no relationship between COVID-19 infection severity and smoking and others that did. Three systematic reviews/meta-analyses reported patients were predominantly middle-aged men (*10*) (*19*) (*40*), and 2 systematic reviews/meta-analyses reported the prevalence of hypertension and diabetes to be around 16% and 8%, respectively (*17*) (*20*). Fever (>80%) and cough (~60%) were the most common symptoms in 2 systemic reviews and meta-analyses (*10*) (*19*) (*40*). Hypertension and diabetes were reported to be associated with more severe disease in several systematic reviews/meta-analyses and ARDS was reported as complication in 9.4% to ~30% of COVID-19 infections. COVID-19 mortality varied anywhere from 3.2 to 14% (*10*) (*20*) (*39*). In pregnant women with COVID-19 infection, the caesarean section rate was >80% and 2 systematic reviews/meta-analyses showed no vertical transmission. Children with COVID-19 presented with mild to moderate symptoms and 2 systematic reviews/meta-analyses reported no deaths in children aged 0 to 9 years **(Table 2)**.

**Table 2.**
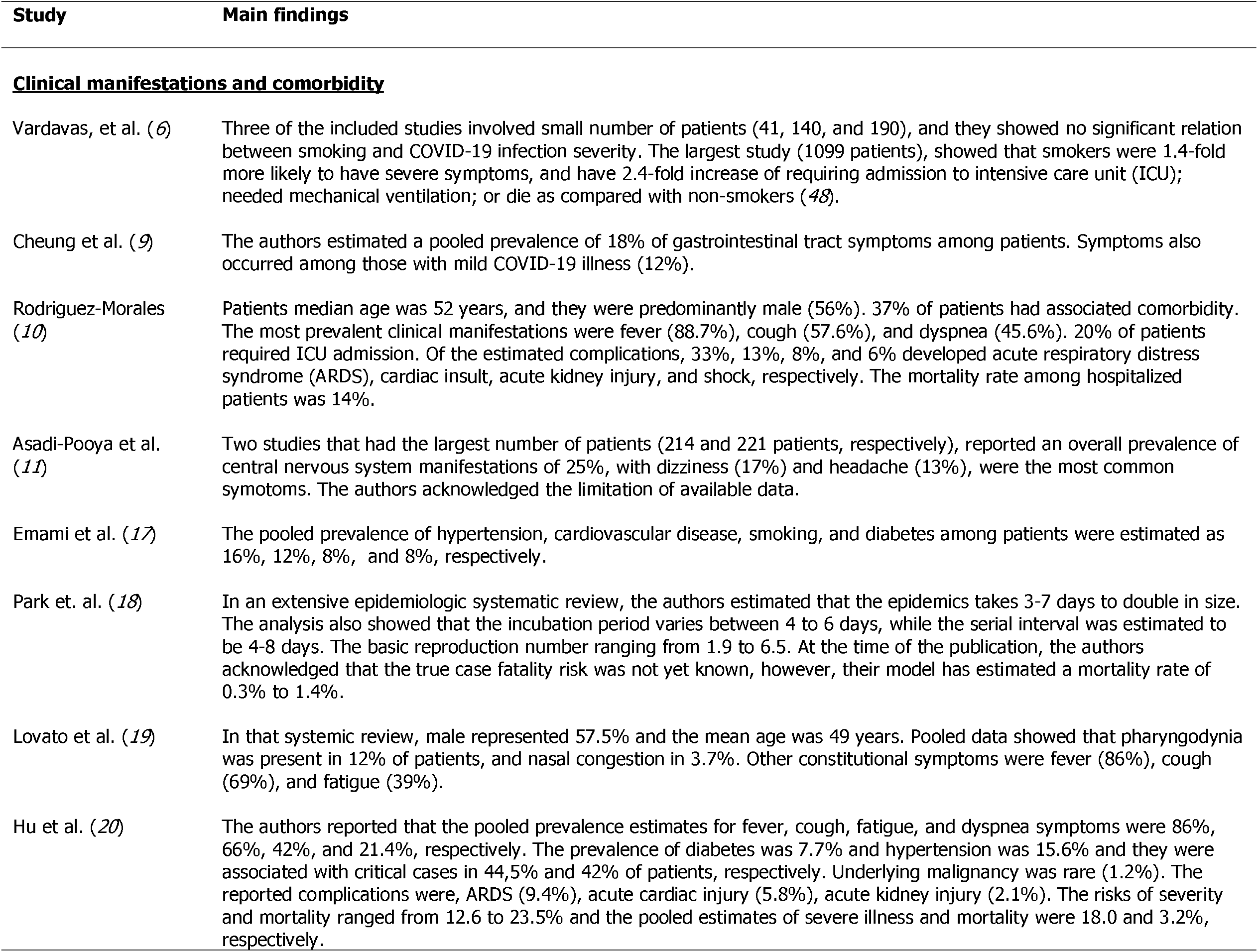
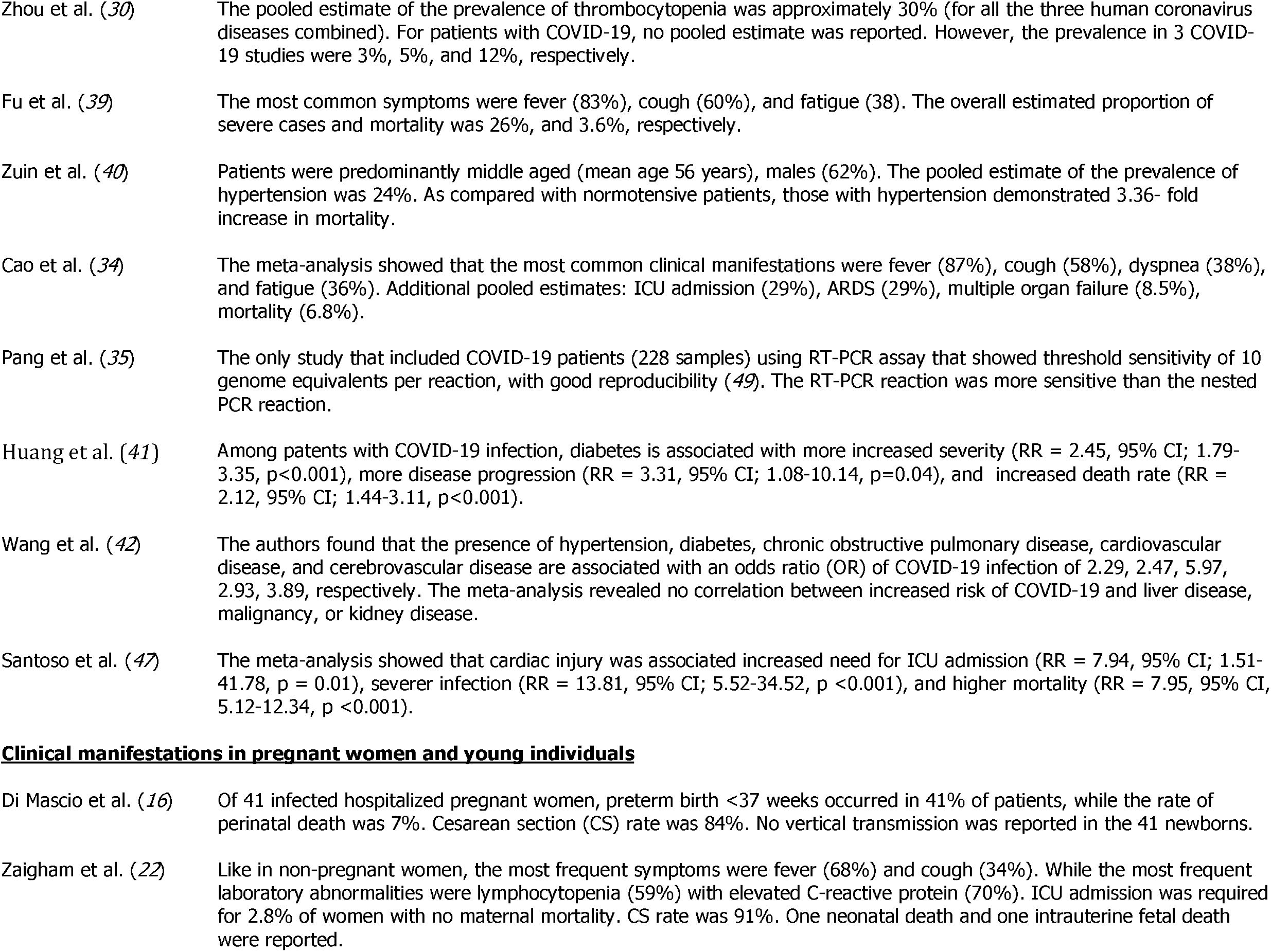
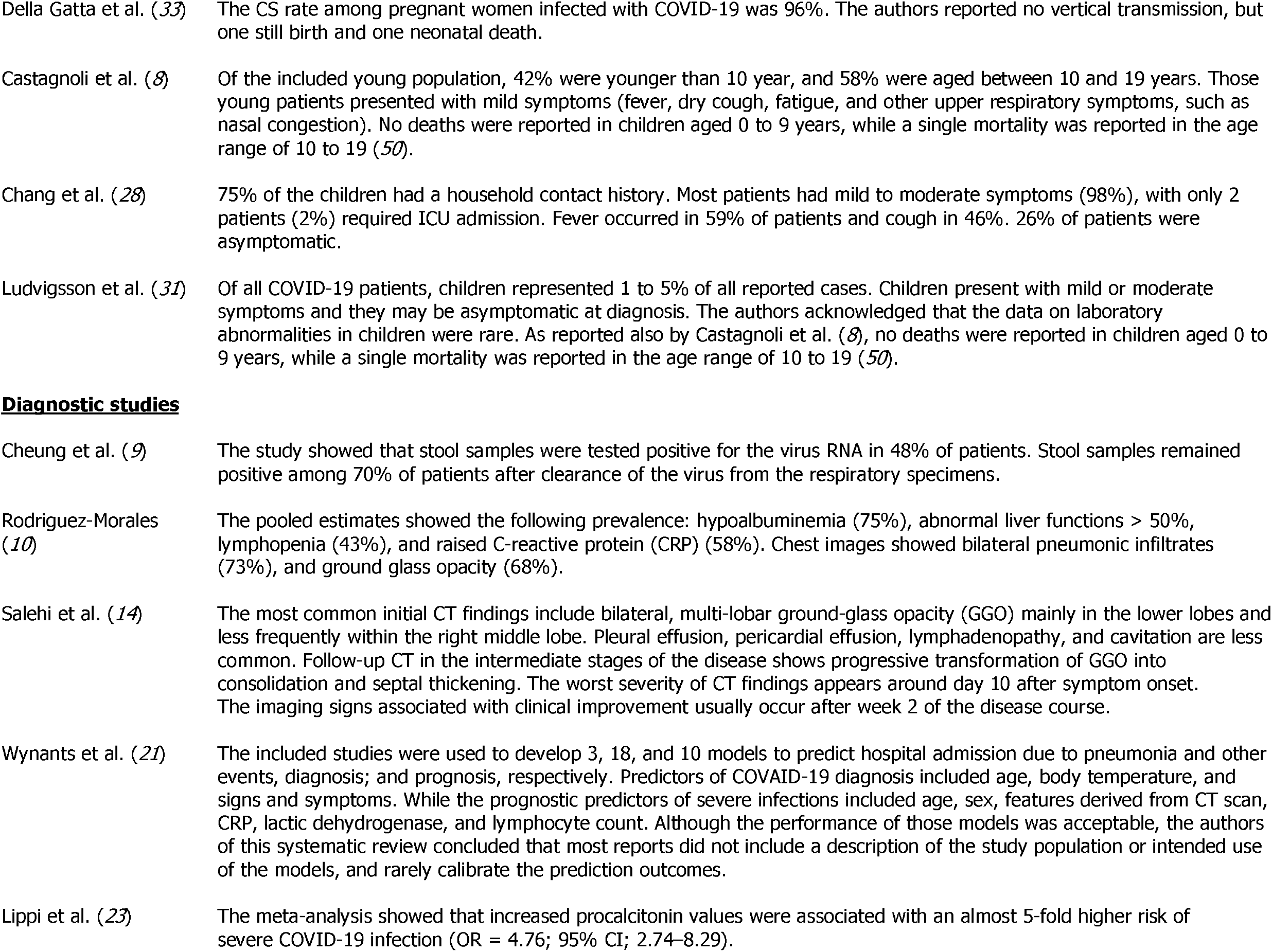
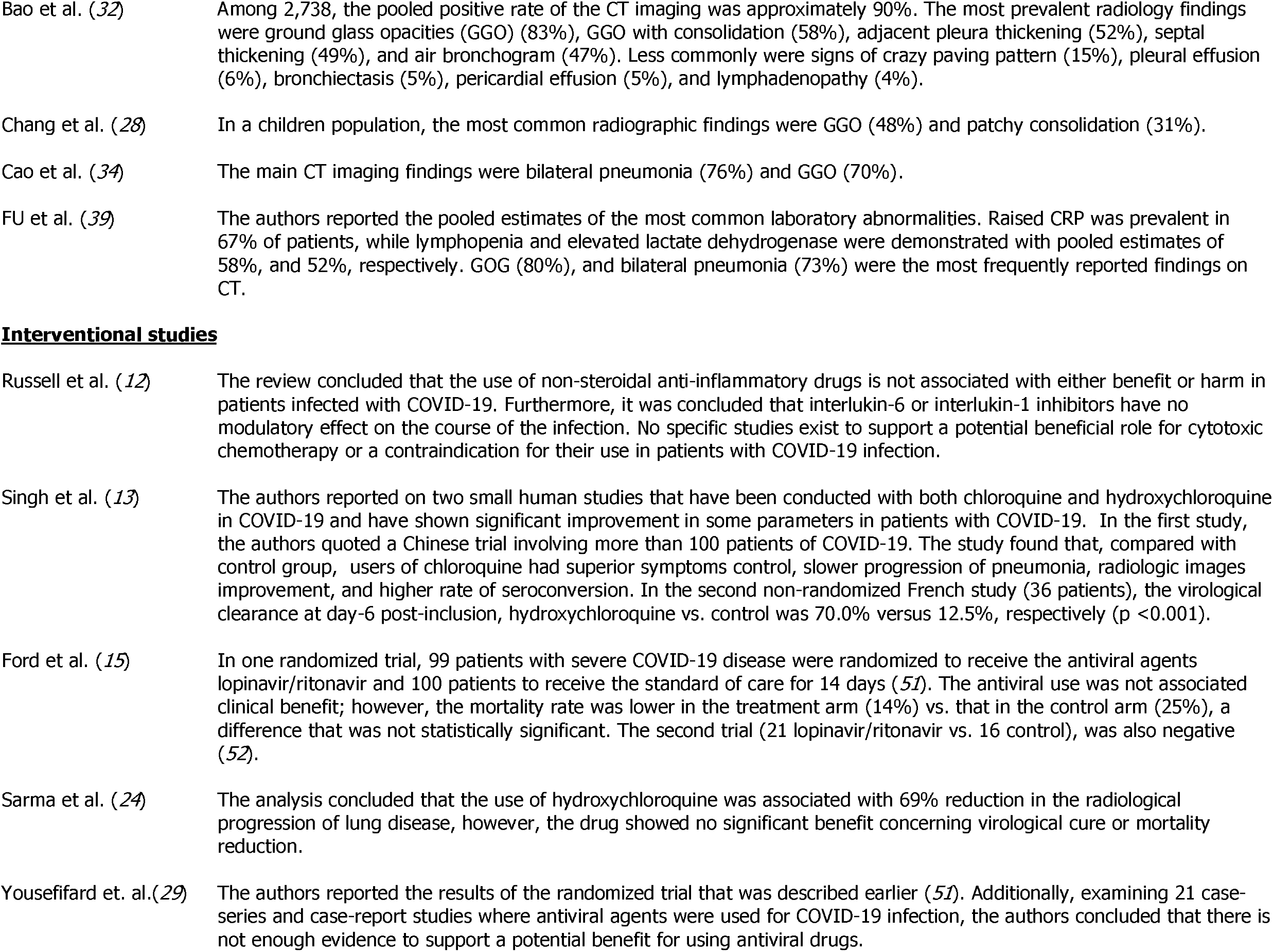
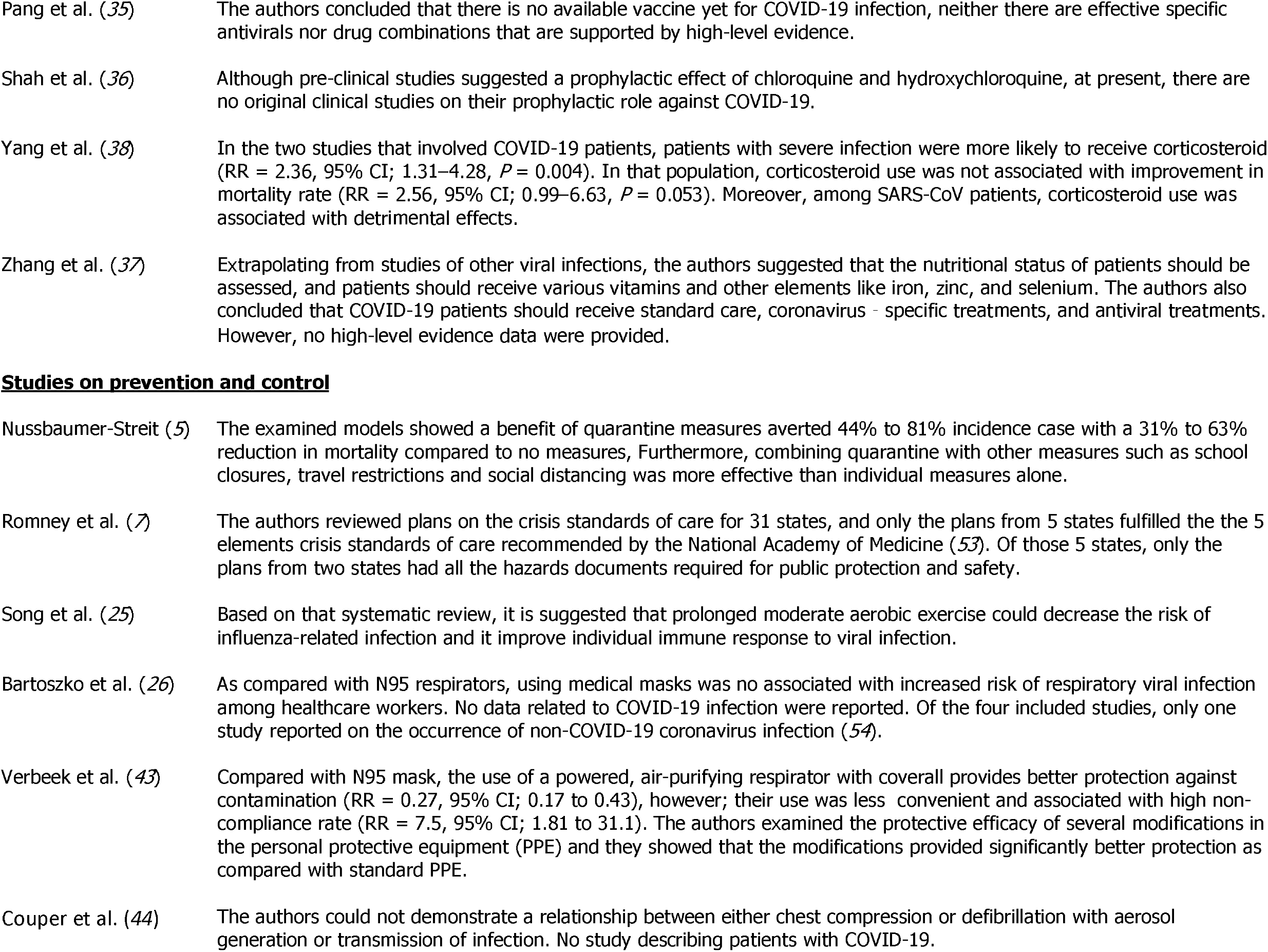
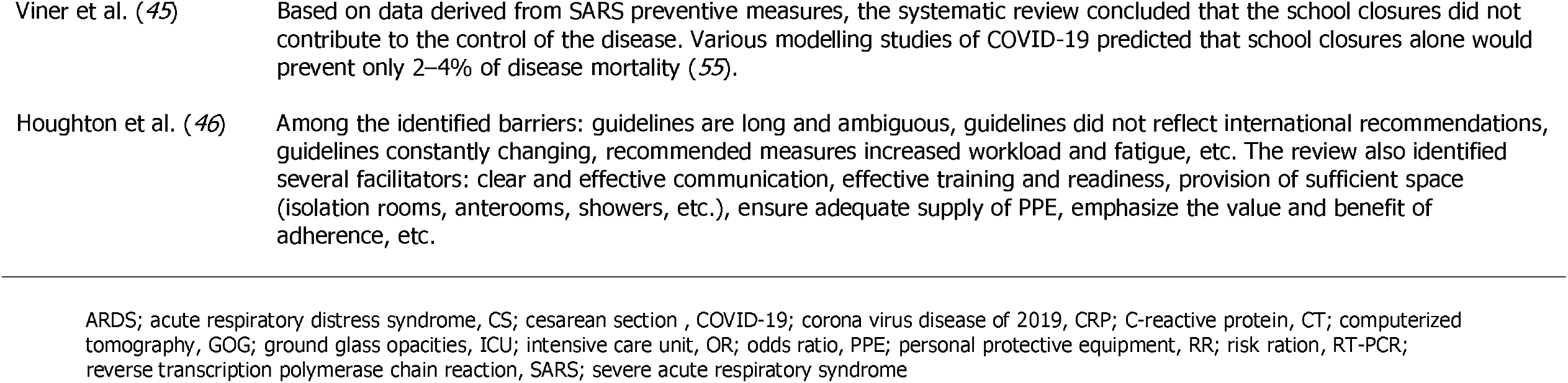
Main findings of the 43 included studies.

| Study | Main findings |
| --- | --- |
| **<u>Clinical manifestations and comorbidity</u>** |  |
| Vardavas, et al. (6) | Three of the included studies involved small number of patients (41, 140, and 190), and they showed no significant relation between smoking and COVID-19 infection severity. The largest study (1099 patients), showed that smokers were 1.4-fold more likely to have severe symptoms, and have 2.4-fold increase of requiring admission to intensive care unit (ICU); needed mechanical ventilation; or die as compared with non-smokers (48). |
| Cheung et al. (9) | The authors estimated a pooled prevalence of 18% of gastrointestinal tract symptoms among patients. Symptoms also occurred among those with mild COVID-19 illness (12%). |
| Rodriguez-Morales (10) | Patients median age was 52 years, and they were predominantly male (56%). 37% of patients had associated comorbidity. The most prevalent clinical manifestations were fever (88.7%), cough (57.6%), and dyspnea (45.6%). 20% of patients required ICU admission. Of the estimated complications, 33%, 13%, 8%, and 6% developed acute respiratory distress syndrome (ARDS), cardiac insult, acute kidney injury, and shock, respectively. The mortality rate among hospitalized patients was 14%. |
| Asadi-Pooya et al. (11) | Two studies that had the largest number of patients (214 and 221 patients, respectively), reported an overall prevalence of central nervous system manifestations of 25%, with dizziness (17%) and headache (13%), were the most common symptoms. The authors acknowledged the limitation of available data. |
| Emami et al. (17) | The pooled prevalence of hypertension, cardiovascular disease, smoking, and diabetes among patients were estimated as 16%, 12%, 8%, and 8%, respectively. |
| Park et. al. (18) | In an extensive epidemiologic systematic review, the authors estimated that the epidemics takes 3-7 days to double in size. The analysis also showed that the incubation period varies between 4 to 6 days, while the serial interval was estimated to be 4-8 days. The basic reproduction number ranging from 1.9 to 6.5. At the time of the publication, the authors acknowledged that the true case fatality risk was not yet known, however, their model has estimated a mortality rate of 0.3% to 1.4%. |
| Lovato et al. (19) | In that systemic review, male represented 57.5% and the mean age was 49 years. Pooled data showed that pharyngodynia was present in 12% of patients, and nasal congestion in 3.7%. Other constitutional symptoms were fever (86%), cough (69%), and fatigue (39%). |
| Hu et al. (20) | The authors reported that the pooled prevalence estimates for fever, cough, fatigue, and dyspnea symptoms were 86%, 66%, 42%, and 21.4%, respectively. The prevalence of diabetes was 7.7% and hypertension was 15.6% and they were associated with critical cases in 44.5% and 42% of patients, respectively. Underlying malignancy was rare (1.2%). The reported complications were, ARDS (9.4%), acute cardiac injury (5.8%), acute kidney injury (2.1%). The risks of severity and mortality ranged from 12.6 to 23.5% and the pooled estimates of severe illness and mortality were 18.0 and 3.2%, respectively. |

**Table 2** Cont.
| Study | Main findings |
| --- | --- |
| Zhou et al. (30) | The pooled estimate of the prevalence of thrombocytopenia was approximately 30% (for all the three human coronavirus diseases combined). For patients with COVID-19, no pooled estimate was reported. However, the prevalence in 3 COVID-19 studies were 3%, 5%, and 12%, respectively. |
| Fu et al. (39) | The most common symptoms were fever (83%), cough (60%), and fatigue (38). The overall estimated proportion of severe cases and mortality was 26%, and 3.6%, respectively. |
| Zuin et al. (40) | Patients were predominantly middle aged (mean age 56 years), males (62%). The pooled estimate of the prevalence of hypertension was 24%. As compared with normotensive patients, those with hypertension demonstrated 3.36- fold increase in mortality. |
| Cao et al. (34) | The meta-analysis showed that the most common clinical manifestations were fever (87%), cough (58%), dyspnea (38%), and fatigue (36%). Additional pooled estimates: ICU admission (29%), ARDS (29%), multiple organ failure (8.5%), mortality (6.8%). |
| Pang et al. (35) | The only study that included COVID-19 patients (228 samples) using RT-PCR assay that showed threshold sensitivity of 10 genome equivalents per reaction, with good reproducibility (49). The RT-PCR reaction was more sensitive than the nested PCR reaction. |
| Huang et al. (41) | Among patents with COVID-19 infection, diabetes is associated with more increased severity (RR = 2.45, 95% CI; 1.79-3.35, $p < 0.001$), more disease progression (RR = 3.31, 95% CI; 1.08-10.14, $p = 0.04$), and increased death rate (RR = 2.12, 95% CI; 1.44-3.11, $p < 0.001$). |
| Wang et al. (42) | The authors found that the presence of hypertension, diabetes, chronic obstructive pulmonary disease, cardiovascular disease, and cerebrovascular disease are associated with an odds ratio (OR) of COVID-19 infection of 2.29, 2.47, 5.97, 2.93, 3.89, respectively. The meta-analysis revealed no correlation between increased risk of COVID-19 and liver disease, malignancy, or kidney disease. |
| Santoso et al. (47) | The meta-analysis showed that cardiac injury was associated increased need for ICU admission (RR = 7.94, 95% CI; 1.51-41.78, $p = 0.01$), severer infection (RR = 13.81, 95% CI; 5.52-34.52, $p < 0.001$), and higher mortality (RR = 7.95, 95% CI, 5.12-12.34, $p < 0.001$). |
| **Clinical manifestations in pregnant women and young individuals** |  |
| Di Mascio et al. (16) | Of 41 infected hospitalized pregnant women, preterm birth <37 weeks occurred in 41% of patients, while the rate of perinatal death was 7%. Cesarean section (CS) rate was 84%. No vertical transmission was reported in the 41 newborns. |
| Zaigham et al. (22) | Like in non-pregnant women, the most frequent symptoms were fever (68%) and cough (34%). While the most frequent laboratory abnormalities were lymphocytopenia (59%) with elevated C-reactive protein (70%). ICU admission was required for 2.8% of women with no maternal mortality. CS rate was 91%. One neonatal death and one intrauterine fetal death were reported. |

**Table 2** Cont.
| Study | Main findings |
| --- | --- |
| Della Gatta et al. (33) | The CS rate among pregnant women infected with COVID-19 was 96%. The authors reported no vertical transmission, but one still birth and one neonatal death. |
| Castagnoli et al. (8) | Of the included young population, 42% were younger than 10 year, and 58% were aged between 10 and 19 years. Those young patients presented with mild symptoms (fever, dry cough, fatigue, and other upper respiratory symptoms, such as nasal congestion). No deaths were reported in children aged 0 to 9 years, while a single mortality was reported in the age range of 10 to 19 (50). |
| Chang et al. (28) | 75% of the children had a household contact history. Most patients had mild to moderate symptoms (98%), with only 2 patients (2%) required ICU admission. Fever occurred in 59% of patients and cough in 46%. 26% of patients were asymptomatic. |
| Ludvigsson et al. (31) | Of all COVID-19 patients, children represented 1 to 5% of all reported cases. Children present with mild or moderate symptoms and they may be asymptomatic at diagnosis. The authors acknowledged that the data on laboratory abnormalities in children were rare. As reported also by Castagnoli et al. (8), no deaths were reported in children aged 0 to 9 years, while a single mortality was reported in the age range of 10 to 19 (50). |
| **Diagnostic studies** |  |
| Cheung et al. (9) | The study showed that stool samples were tested positive for the virus RNA in 48% of patients. Stool samples remained positive among 70% of patients after clearance of the virus from the respiratory specimens. |
| Rodriguez-Morales (10) | The pooled estimates showed the following prevalence: hypoalbuminemia (75%), abnormal liver functions > 50%, lymphopenia (43%), and raised C-reactive protein (CRP) (58%). Chest images showed bilateral pneumonic infiltrates (73%), and ground glass opacity (68%). |
| Salehi et al. (14) | The most common initial CT findings include bilateral, multi-lobar ground-glass opacity (GGO) mainly in the lower lobes and less frequently within the right middle lobe. Pleural effusion, pericardial effusion, lymphadenopathy, and cavitation are less common. Follow-up CT in the intermediate stages of the disease shows progressive transformation of GGO into consolidation and septal thickening. The worst severity of CT findings appears around day 10 after symptom onset. The imaging signs associated with clinical improvement usually occur after week 2 of the disease course. |
| Wynants et al. (21) | The included studies were used to develop 3, 18, and 10 models to predict hospital admission due to pneumonia and other events, diagnosis; and prognosis, respectively. Predictors of COVID-19 diagnosis included age, body temperature, and signs and symptoms. While the prognostic predictors of severe infections included age, sex, features derived from CT scan, CRP, lactic dehydrogenase, and lymphocyte count. Although the performance of those models was acceptable, the authors of this systematic review concluded that most reports did not include a description of the study population or intended use of the models, and rarely calibrate the prediction outcomes. |
| Lippi et al. (23) | The meta-analysis showed that increased procalcitonin values were associated with an almost 5-fold higher risk of severe COVID-19 infection (OR = 4.76; 95% CI; 2.74–8.29). |

**Table 2** Cont.
| Study | Main findings |
| --- | --- |
| Bao et al. (32) | Among 2,738, the pooled positive rate of the CT imaging was approximately 90%. The most prevalent radiology findings were ground glass opacities (GGO) (83%), GGO with consolidation (58%), adjacent pleura thickening (52%), septal thickening (49%), and air bronchogram (47%). Less commonly were signs of crazy paving pattern (15%), pleural effusion (6%), bronchiectasis (5%), pericardial effusion (5%), and lymphadenopathy (4%). |
| Chang et al. (28) | In a children population, the most common radiographic findings were GGO (48%) and patchy consolidation (31%). |
| Cao et al. (34) | The main CT imaging findings were bilateral pneumonia (76%) and GGO (70%). |
| FU et al. (39) | The authors reported the pooled estimates of the most common laboratory abnormalities. Raised CRP was prevalent in 67% of patients, while lymphopenia and elevated lactate dehydrogenase were demonstrated with pooled estimates of 58%, and 52%, respectively. GOG (80%), and bilateral pneumonia (73%) were the most frequently reported findings on CT. |
| **Interventional studies** |  |
| Russell et al. (12) | The review concluded that the use of non-steroidal anti-inflammatory drugs is not associated with either benefit or harm in patients infected with COVID-19. Furthermore, it was concluded that interleukin-6 or interleukin-1 inhibitors have no modulatory effect on the course of the infection. No specific studies exist to support a potential beneficial role for cytotoxic chemotherapy or a contraindication for their use in patients with COVID-19 infection. |
| Singh et al. (13) | The authors reported on two small human studies that have been conducted with both chloroquine and hydroxychloroquine in COVID-19 and have shown significant improvement in some parameters in patients with COVID-19. In the first study, the authors quoted a Chinese trial involving more than 100 patients of COVID-19. The study found that, compared with control group, users of chloroquine had superior symptoms control, slower progression of pneumonia, radiologic images improvement, and higher rate of seroconversion. In the second non-randomized French study (36 patients), the virological clearance at day-6 post-inclusion, hydroxychloroquine vs. control was 70.0% versus 12.5%, respectively ($p < 0.001$). |
| Ford et al. (15) | In one randomized trial, 99 patients with severe COVID-19 disease were randomized to receive the antiviral agents lopinavir/ritonavir and 100 patients to receive the standard of care for 14 days (51). The antiviral use was not associated clinical benefit; however, the mortality rate was lower in the treatment arm (14%) vs. that in the control arm (25%), a difference that was not statistically significant. The second trial (21 lopinavir/ritonavir vs. 16 control), was also negative (52). |
| Sarma et al. (24) | The analysis concluded that the use of hydroxychloroquine was associated with 69% reduction in the radiological progression of lung disease, however, the drug showed no significant benefit concerning virological cure or mortality reduction. |
| Yousefifard et. al.(29) | The authors reported the results of the randomized trial that was described earlier (51). Additionally, examining 21 case-series and case-report studies where antiviral agents were used for COVID-19 infection, the authors concluded that there is not enough evidence to support a potential benefit for using antiviral drugs. |

**Table 2** Cont.
| Study | Main findings |
| --- | --- |
| Pang et al. (35) | The authors concluded that there is no available vaccine yet for COVID-19 infection, neither there are effective specific antivirals nor drug combinations that are supported by high-level evidence. |
| Shah et al. (36) | Although pre-clinical studies suggested a prophylactic effect of chloroquine and hydroxychloroquine, at present, there are no original clinical studies on their prophylactic role against COVID-19. |
| Yang et al. (38) | In the two studies that involved COVID-19 patients, patients with severe infection were more likely to receive corticosteroid (RR = 2.36, 95% CI; 1.31–4.28, $P = 0.004$). In that population, corticosteroid use was not associated with improvement in mortality rate (RR = 2.56, 95% CI; 0.99–6.63, $P = 0.053$). Moreover, among SARS-CoV patients, corticosteroid use was associated with detrimental effects. |
| Zhang et al. (37) | Extrapolating from studies of other viral infections, the authors suggested that the nutritional status of patients should be assessed, and patients should receive various vitamins and other elements like iron, zinc, and selenium. The authors also concluded that COVID-19 patients should receive standard care, coronavirus - specific treatments, and antiviral treatments. However, no high-level evidence data were provided. |
| **Studies on prevention and control** |  |
| Nussbaumer-Streit (5) | The examined models showed a benefit of quarantine measures averted 44% to 81% incidence case with a 31% to 63% reduction in mortality compared to no measures, Furthermore, combining quarantine with other measures such as school closures, travel restrictions and social distancing was more effective than individual measures alone. |
| Romney et al. (7) | The authors reviewed plans on the crisis standards of care for 31 states, and only the plans from 5 states fulfilled the the 5 elements crisis standards of care recommended by the National Academy of Medicine (53). Of those 5 states, only the plans from two states had all the hazards documents required for public protection and safety. |
| Song et al. (25) | Based on that systematic review, it is suggested that prolonged moderate aerobic exercise could decrease the risk of influenza-related infection and it improve individual immune response to viral infection. |
| Bartoszko et al. (26) | As compared with N95 respirators, using medical masks was no associated with increased risk of respiratory viral infection among healthcare workers. No data related to COVID-19 infection were reported. Of the four included studies, only one study reported on the occurrence of non-COVID-19 coronavirus infection (54). |
| Verbeek et al. (43) | Compared with N95 mask, the use of a powered, air-purifying respirator with coverall provides better protection against contamination (RR = 0.27, 95% CI; 0.17 to 0.43), however; their use was less convenient and associated with high non-compliance rate (RR = 7.5, 95% CI; 1.81 to 31.1). The authors examined the protective efficacy of several modifications in the personal protective equipment (PPE) and they showed that the modifications provided significantly better protection as compared with standard PPE. |
| Couper et al. (44) | The authors could not demonstrate a relationship between either chest compression or defibrillation with aerosol generation or transmission of infection. No study describing patients with COVID-19. |

**Table 2** Cont.
| Study | Main findings |
| --- | --- |
| Viner et al. (45) | Based on data derived from SARS preventive measures, the systematic review concluded that the school closures did not contribute to the control of the disease. Various modelling studies of COVID-19 predicted that school closures alone would prevent only 2–4% of disease mortality (55). |
| Houghton et al. (46) | Among the identified barriers: guidelines are long and ambiguous, guidelines did not reflect international recommendations, guidelines constantly changing, recommended measures increased workload and fatigue, etc. The review also identified several facilitators: clear and effective communication, effective training and readiness, provision of sufficient space (isolation rooms, anterooms, showers, etc.), ensure adequate supply of PPE, emphasize the value and benefit of adherence, etc. |

ARDS; acute respiratory distress syndrome, CS; cesarean section, COVID-19; corona virus disease of 2019, CRP; C-reactive protein, CT; computerized tomography, GOG; ground glass opacities, ICU; intensive care unit, OR; odds ratio, PPE; personal protective equipment, RR; risk ratio, RT-PCR; reverse transcription polymerase chain reaction, SARS; severe acute respiratory syndrome

The most frequent chest CT findings were GGO and bilateral infiltrates in several systematic reviews/meta-analyses and 1 meta-analysis showed increased procalcitonin concentrations were associated with higher risk for severe COVID-19 infections (*23*). With regards to medication interventions, 3 systematic reviews/meta-analyses reported no benefit for using antiviral drugs (*15*) (*29*) (*35*). One systematic review concluded neither benefit or harm with use on non-steroidal anti-inflammatory drugs and that interleukin-1 and −6 have no modulatory effects on the course of COVID-19 infections (*12*). Systematic reviews/meta-analyses on the effects of chloroquine and hydroxychloroquine showed varying results. One systematic review noted superior symptom control and improvement in radiographic imaging in one study and superior virological clearance compared to control in another study (*13*). Another systematic review/meta-analysis also showed reduction in radiologic progression of disease, but no significant benefit regarding virological cure or mortality reduction (*24*) **(Table 2)**.

Systematic reviews/meta-analyses on prevention and control of COVID-19 provided occasionally conflicting results as well. For example, one review concluded that school closures did not contribute to control of SARS and modeling for COVID-19 predicted school closures would only prevent 2-4% disease mortality (*45*); while another review that combined COVID-19 and non-COVID-19 studies reported that quarantine combined with school closures and other measures was effective (5). One systematic review/meta-analysis examining medical masks vs N95 respirators for preventing COVID-19 in healthcare workers did not include any COVID-19 studies (*26*).

## Discussion

Of the 7850 potentially relevant articles, 43 systematic reviews/meta-analyses met our inclusion/exclusion criteria were included **(Figure 1)**. The various studies included addressed clinical manifestations and associated comorbidity, clinical manifestations in pregnant women and younger individuals, diagnostic data, various interventions, and prevention and control of COVID-19. As noted, we did not intent to evaluate the quality of published studies but noted occasionally conflicting data, large ranges for COVID-19 associated acute respiratory distress syndrome (ARDS) and mortality estimates, systematic reviews/meta-analyses that included non-COVID-19 trials, similarities for comorbidities increasing disease severity such as diabetes and hypertension, and predominance of middle-aged male patients with COVID-19 infections.

**Figure 1.**
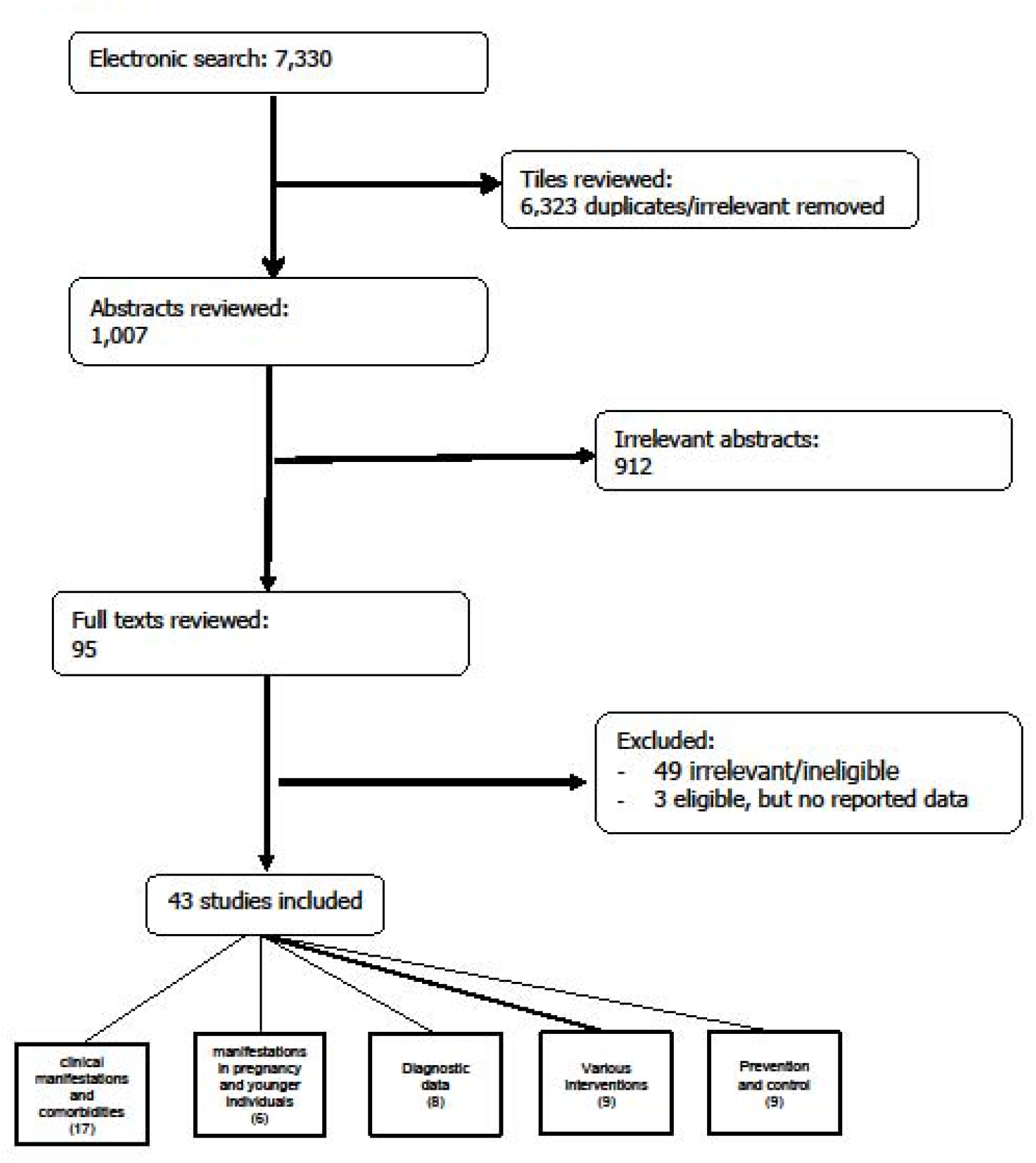
The flowchart of the literature search and the selection of the 43 studies. Some studies have addressed more than one question.

Following the outbreak of COVID-19 in Wuhan in the Hubei Province of China in December 2019 (1) and the subsequent declaration by the WHO of a pandemic that has caused alarming mortality and disruption of the world economy affecting several millions of people worldwide, there was need for rapid dissemination of information regarding this novel infection as more clinicians and scientists dealt with the reality of COVID-19. Technology and social media played a large part in the sharing of clinical information and experiences with disease management-Chinese clinicians communicated their experiences with European and American clinicians via virtual conferences, for example, because the virus had affected their community first. The information disseminated this way was based on clinical experiences and the patients seen in their hospitals who may differ in comorbidities from patients seen in Europe and North America, however, the dissemination of information was critical to guide clinicians.

We also witnessed a flood of published manuscripts in several journals, including highly impactful ones. Some published studies included small numbers of patients and it is likely the studies were published to share as much information as possible given the devastating effects of COVID-19 on millions globally. As more patients became ill with COVID-19, as hospitals became overwhelmed with sick patients with various ranges of disease severity and ran out of beds and ventilators, as personal protective equipment was challenging to secure in some countries, as various medications were attempted for prophylactic use and treatment, as financial institutions suffered major loses, as weaknesses in healthcare systems were exposed, and most important, as millions of lives were lost, the internet served as an integral space to share both accurate and inaccurate information; including published and retracted studies.

There were common clinical manifestations and comorbidities in the systematic reviews/meta-analyses we included. Notably, patients were predominantly middle-aged men and comorbidities such a diabetes, hypertension, cardiovascular disease, and smoking were prevalent and associated with more severe disease. The most common clinical manifestations were fever, cough, and dyspnea. Rates of ARDS and mortality varied between studies. In pregnant women with COVID-19 infection, the caesarean section rate was >80% and children with COVID-19 presented with mild to moderate symptoms and no deaths were reported in children aged 0 to 9 years **(Table 2)**. The most frequent chest CT findings were GGO and bilateral infiltrates. With regards to medication interventions, no benefit for using antiviral drugs was found, no benefit or harm with use of non-steroidal anti-inflammatory drugs was found, and no benefit with interleukin-1 and −6 was found. Radiographic improvement and better symptom control were reported with chloroquine and hydroxychloroquine. Finally, quarantining, social distancing, and school closures may contribute to reduction of disease spread and may be more effective if used in combination **(Table 2)**.

The ability to rapidly disseminate information via the rapid review process some journals implemented is a double-edged sword. It remains of utmost important to maintain the integrity of published science while keeping in mind the devastation this COVID-19 has caused and the desperate need for a cure, medications that will lessen disease severity, and/or a vaccine. Additionally, care must be taken to distinguish between anecdotal experiences and data from randomized controlled trials- the standard clinical trials based on which treatment guidelines are usually created.

As more data is collected from patients infected with COVID-19 all over the world, more studies undoubtedly will be published. As clinicians, scientists, and members of society, we will look forward to the sharing of information. Scientists, reviewers, editors, and journals alike must work to maintain the integrity of the science shared. The internet, social media, the ability to hold virtual conferences across the globe, and the yearning to find a cure will promote dissemination of clinical experiences and science. This pandemic will not only change the way we live, but also encourage the use of non-conventional methods to share science.

## Data Availability

Not applicable

